# A single-nucleus transcriptomics study of alcohol use disorder in the nucleus accumbens

**DOI:** 10.1101/2022.06.16.22272431

**Authors:** Edwin J.C.G. van den Oord, Lin Y Xie, Min Zhao, Karolina A. Aberg, Shaunna L. Clark

**Author notes:** Correspondence: Edwin J.C.G. van den Oord, Ph.D., McGuire Hall, Room 216A, 1112 East Clay Street, Richmond, P. O. Box 980533, VA 23298-0581.

## Abstract

Alcohol use disorder (AUD) is a significant public health problem. Gene expression studies offer promising opportunities to better understand the underlying pathogenic processes. As cell-types differ in their function, gene expression profiles will typically vary across cell-types. When studying bulk tissue, failure to account for this cellular diversity has a detrimental impact on the ability to detect disease associations. We therefore assayed the transcriptomes of 32,531 individual nuclei extracted from the nucleus accumbens (NAc) of 9 donors with AUD and 9 controls. Our study identified 17 clearly delineated cell-types. We detected 26 transcriptome-wide significant association signals (q-value<0.1) that mainly involved medium spiny neurons with both D1-type and D2-type dopamine receptors, microglia and oligodendrocytes. A significantly higher number of findings than expected by chance replicated in an existing single nucleus gene expression study of alcohol dependence in the pre-frontal cortex (enrichment ratio 1.91, P value 0.019). The alcohol related genes and pathways detected for each cell-type were consistent with the functions of those cell-types reported in the literature. Thus, for the neurons we observed alcohol related neurodegeneration, disruption of circadian rhythms, alterations in glucose metabolism, and changes in synaptic plasticity. For microglia we found neuroinflammation and immune-related processes and for oligodendrocytes disruptions in myelination. This identification of the specific cell-types from which the association signals originate is key for designing proper follow-up experiments and, eventually, for developing new and targeted clinical interventions.

## INTRODUCTION

Alcohol use disorder (AUD) is characterized by an impaired ability to stop or control alcohol use despite adverse social, occupational, or health consequences. Gene expression studies offer promising opportunities to better understand the underlying pathogenic processes. As cells differ in their function, gene expression profiles will typically differ across cell-types. When studying bulk tissue, failure to account for the cellular diversity has a detrimental impact on the ability to detect disease associations(1). For example, case-control differences will be “diluted” if they involve only one cell-type, may cancel out if the differences are of opposite signs across cell-types, and may be undetectable if the differences involve low abundance cells. Furthermore, identifying the specific cell-types from which the association signals originate is key to formulating refined hypotheses of AUD pathology, designing proper follow-up experiments and, eventually, developing effective clinical interventions.

With the recent development of single cell/nucleus sequencing technologies (s_c/n_RNA-seq) it has become possible to characterize the expression levels of thousands of individual cells with a single assay thereby allowing cell-type specific gene expression studies on a very refined level. In comparison to whole cells, nuclei are more resistant to mechanical assaults and are less vulnerable to the tissue dissociation process. This makes s_n_-RNA-seq the more suitable option for (frozen) post-mortem brain tissue(2, 3). Only one s_n_RNA-seq study of alcohol has been reported that involved nuclei from the pre-frontal cortex of 3 alcohol-dependent patients and 4 controls(4). The authors assigned nuclei to one of seven known brain cell-types, identified differentially expressed genes, and demonstrated that many of the cell-type specific findings were not present when bulk tissue was considered(4).

In this study we assayed the transcriptomes of 32,531 individual nuclei extracted from the nucleus accumbens (NAc) of 9 human donors with AUD and 9 controls. We chose the NAc due to its central role in the mesolimbic reward pathway, and being a well-studied target for neuromodulatory therapies for AUD(5).

## MATERIALS AND METHODS

The supplemental material provides details on the data generation and analyses.

### Sample description

NAc tissue was obtained through the NIH NeuroBioBank from 9 cases and 9 controls from the Human Brain Collection Core. The cases had a confirmed DSM-IV diagnosis for alcohol dependence. Controls had no previous history of alcohol dependence. With the exception of major depressive disorder (MDD), the donors did not have any neurological or psychiatric condition.

### s_n_RNA-seq data generation, alignment and quality control

Nuclei were isolated using a further optimized protocol specifically developed for frozen human biobanked brain tissue(6). Isolated nuclei were partitioned using the Chromium system from 10X Genomics. Following paired-end library preparation each sample was sequenced on a NovaSeq 6000. The cellranger(7) software was used for de-multiplexing the expression information from each nucleus, aligning the reads to GRCh38, and creating a matrix with counts of the number of unique molecules for each gene detected in each nucleus. Nuclei contain a relatively large fraction of unspliced pre-mRNA molecules and such molecules are particularly abundant in brain tissue(8). As pre-mRNA transcripts can generate intronic reads(9), we aligned reads using a gene transfer format file that allowed intronic alignments.

We quality controlled (QC’ed) nuclei and genes. This included removing low-quality nuclei or empty droplets, as indicated by having only a few genes expressed, and removing nuclei multiplets, as indicated by having many genes expressed. We also removed genes with low abundance levels as well as genes that are only observed in a small number of nuclei. This gene QC was performed for each cell-type separately rather than across all cells. This avoids, for instance, that genes that are highly expressed in only one cell-type are eliminated because of low expression levels in all the other cell-types. The data was log-normalized to reduce effects of possible outliers and scaled to have unit variance to avoid that highly-expressed genes dominate the cluster analyses.

### Clustering and labeling of cell-types

Cluster analyses of nuclei were perform in Seurat(10). To improve clustering, these analyses were restricted to only a limited set of genes that exhibit high nucleus-to-nucleus variation(11). There are potentially a large number of subject-level variables (e.g., sex, age) and confounders (e.g., cDNA yield, percentage of reads aligned) that may obscure the separation of clusters. To remove this subject-level variation, prior to clustering we regressed out “dummy” variables that indicated the 18 donors. Furthermore, we regressed out nuclei QC measures (e.g., number of genes per nucleus). A non-linear dimensional reduction technique (tSNE) was used to visualize the clusters. The clusters were labelled using known cell-type specific gene expression markers.

### Identifying differentially expressed genes (DEGs)

The (fixed) effect of case-control status was tested using mixed models (R nlme package) that included a random intercept to account for the dependency in the data that results from assaying many nuclei from the same donors. Covariates in the analyses were (i) Chromium batch plus the nuclei QC measures (ii) sex and race, and (iii) MDD status. In addition, we regressed out principal components (PCs) to capture any remaining unmeasured confounders. The principal component analysis was performed after regressing out the covariates as well as indicator variables for the cell clusters to avoid that the PCs captured biological variation between nuclei. We controlled the false discovery rate at the 0.1 level as that provides a good balance between the competing aims of finding true associations and avoiding false discoveries(12).

## RESULTS

### Sample and assay related statistics

Table 1 shows that were no significant differences between cases and controls on key donor and assay related statistics (Table S2 provides a full set of statistics). The exception was MDD, which was a covariate in the analyses. Samples were sequenced at a relatively high depth of 241,497 high-quality reads per nucleus to ensure low expressed genes could be detected. The total number of nuclei was 32,531 of which 28,494 (87.6%) remained after QC.

**Table 1.** Descriptive statistics

|  | Controls (N=9) |  | AUD cases (N=9) |  | P value* |
| --- | --- | --- | --- | --- | --- |
|  | Mean/<br>Count | SD/<br>% | Mean/<br>Count | SD/<br>% |  |
| Sample related statistics |  |  |  |  |  |
| Age (years) | 44.2 | 7.4 | 44.9 | 8.3 | 0.86 |
| Male | 6 | 67% | 7 | 78% | 0.08 |
| European American** | 6 | 67% | 8 | 89% | 0.33 |
| PMI (hours) | 31.9 | 14.9 | 31.2 | 7.8 | 0.89 |
| MDD** | 0 | 0% | 4 | 44% | *** |
| Assay related statistics |  |  |  |  |  |
| % Reads mapped | 93.8 | 2 | 93.4 | 3.2 | 0.71 |
| Q30 bases in reads | 92.7 | 0.5 | 92.7 | 0.5 | 1 |
| % Reads in nuclei | 81.4 | 5.6 | 81.6 | 6.2 | 0.97 |
| Number of nuclei | 2,060 | 2,390 | 1,556 | 1,138 | 0.58 |
| Mean reads/nucleus | 262,101 | 167,523 | 220,893 | 88,152 | 0.53 |
| Median genes/nucleus | 3,740 | 1,018 | 3,839 | 1,076 | 0.84 |

Note: PMI is post-mortem interval, MDD is major depressive disorder. \* We used a T-test for quantitative variables and Fisher's exact test for categorical variables. \*\* Remaining donors are African American. \*\*\* P value cannot be computed as MDD has no variation in controls.

### Clustering and labeling of cell-types

We identified 17 clusters (Figure 1). Table S3 lists the key discriminating genes as determined by MAST(13). Table S4 provides the full list of gene expression markers used to label the clusters. Standard markers readily identified endothelial cells (END)(14), oligodendrocytes (OLI) and oligodendrocyte precursor cells (OPC)(15), and microglia (MGL)(16). Four subclusters of spiny GABAergic interneurons (INT) were identified. Three subclusters could be labeled based on the expression of somatostatin (INT.SST), parvalbumin (INT.PV), and vasoactive intestinal peptide (INT.VIP)(17). The fourth expressed *DRD1* and D1-like markers but not *DRD2* and D2-like markers(18) and was labeled INT.D1. Astrocytes (ASC) were detected based on standard markers(19). The marker profiles of each of the three subcluster were most highly correlated with their “AST1” marker profile identified in(19) that the authors considered mature astrocytes. One subcluster of astrocytes expressed the MGL marker *P2RY12* and OLI marker MBP (Myelin basic protein) potentially suggesting they may affect myelination (ASC.MYL)(20). The NAc lacks glutamatergic neurons but instead has GABAergic inhibitory medium-sized spiny neurons (MSNs)(21). The biggest subcluster (MSN.D1) expressed *DRD1* (D1) but not *DRD2* (D2) or D2 related markers such as *PENK*. The other subclusters co-expressed D1 and D2. The percentage of mixed D1/D2 MSNs was higher than expected. As mixed MSNs are more often observed in the NAc shell than the core(21), one possible explanation could be that our samples were predominantly from the shell. One of the subclusters (MSN.HTR) uniquely expressed *HTR2C* at high levels, a phenomenon observed previously(22). *ALDH1A1* is an astrocyte marker that can be expressed in certain midbrain dopaminergic neurons(23). The expression of this marker was unique for a single MSN subcluster (MSN.ALDH).

**Figure 1.**
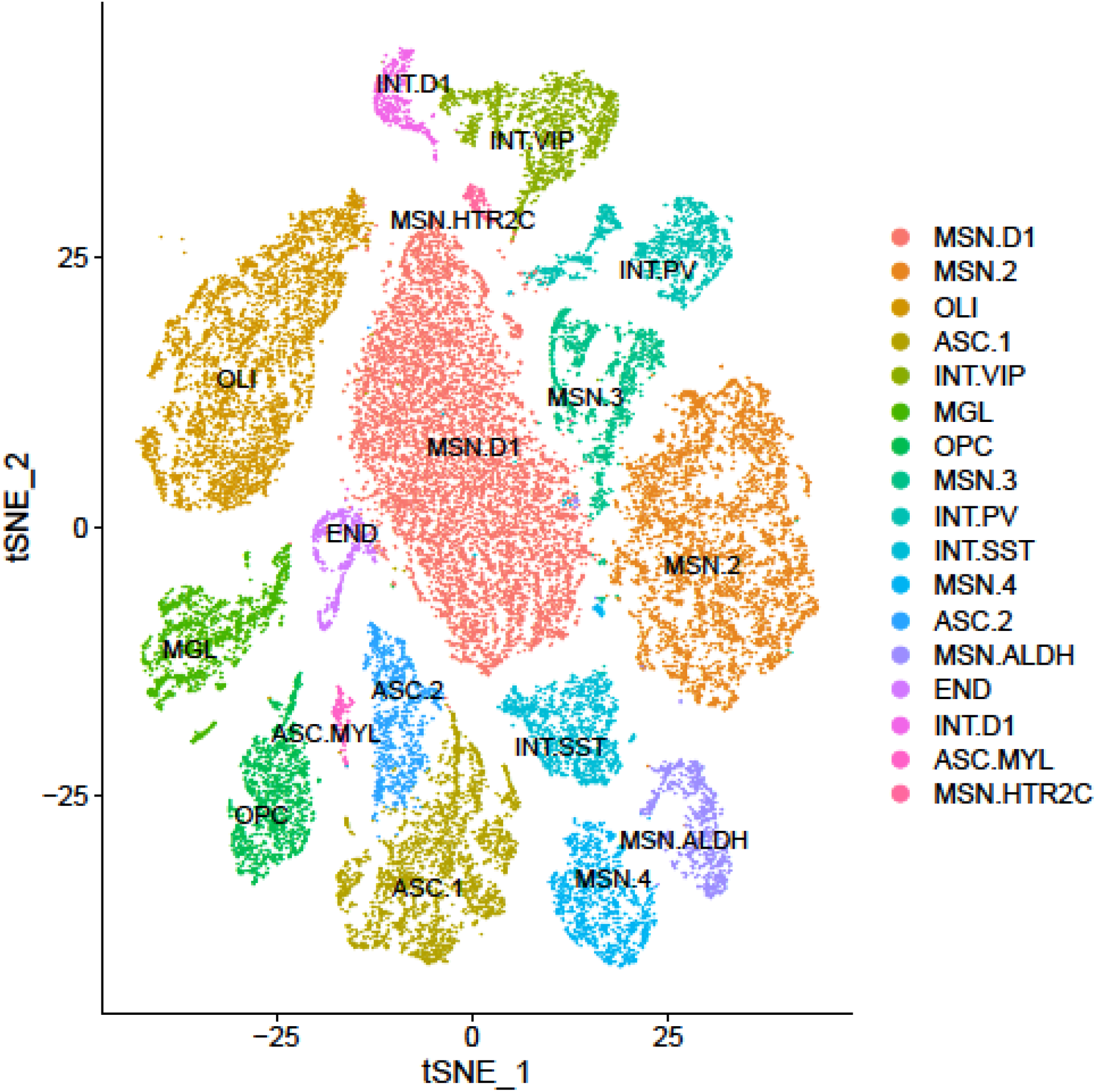
A *t*-distributed stochastic neighbor embedding (t-SNE) plot depicting the nuclei in 17 clusters. The figure shows one cluster of endothelial cells (END), oligodendrocytes (OLI), oligodendrocyte precursor cells (OPC), and microglia (MGL). Three subclusters of astrocytes (ASC) are displayed, one of which are myelin astrocytes (ASC.MYL). The four subclusters of spiny GABAergic interneurons (INT) could be further labeled based on the expression of somatostatin (INT.SST), parvalbumin (INT.PV), and vasoactive intestinal peptide (INT.VIP) and *DRD1* (INT.D1). Five subclusters of GABAergic inhibitory medium-sized spiny neurons (MSNs) are shown three subclusters uniquely expressing *DRD1* (MSN.D1), *HTR2C* (MSN.HTR) and *ALDH1A1* (MSN.ALDH).

### Identifying differentially expressed genes (DEGs)

The total number of genes detected was 36,601. After cell-type specific QC of low abundant genes, on average 4,287 genes were tested per cell-type. The mean/median lambda (the median of the observed test statistics divided by the expected median of the test statistics under the assumed null distribution) across the 17 cell-types was 1.08/1.03. This suggested the absence of test statistic inflation and that our P values were accurate. Analyses were re-run using more liberal QC thresholds. Although this increased the number of significant results, it also increased lambda meaning that findings are more likely false discoveries due to test statistic inflation.

We obtained 26 DEGs (q-value<0.1, Table 2), with 113 genes reaching suggestive significance (q-value<0.25, Table S6). The DEGs included 16 findings for MSN.3, 6 for MGL, 2 for OLI, 1 for OPC, and 1 for ASC.1. Four DEGs involved lncRNA and the rest protein coding genes. P values ranged from 1.17×10^−5^ for *TMEM178B* in OPC to 3.83×10^−4^ *XPO1* in MGL.

**Table 2.** Significant association results after FDR control at the 0.1 level

| Cell | Gene | Chr | Start | End | Type | b | T_stat | P value | Q_value |
| --- | --- | --- | --- | --- | --- | --- | --- | --- | --- |
| ASC.1 | CCDC18.AS1 | 1 | 93261701 | 93346434 | lncRNA | -0.271 | -6.32 | 1.03E-05 | 2.62E-02 |
| MGL | CEP85L | 6 | 118460772 | 118710075 | PCG | 0.392 | 5.84 | 2.53E-05 | 3.02E-02 |
| MGL | SUMF1 | 3 | 3700814 | 4467273 | PCG | 0.397 | 5.11 | 1.06E-04 | 4.46E-02 |
| MGL | TRAF3 | 14 | 102777476 | 102911500 | PCG | -0.397 | -4.97 | 1.41E-04 | 4.46E-02 |
| MGL | CD53 | 1 | 110871188 | 110899922 | PCG | -0.775 | -4.94 | 1.49E-04 | 4.46E-02 |
| MGL | RAPH1 | 2 | 203394345 | 203535335 | PCG | -0.43 | -4.64 | 2.73E-04 | 6.52E-02 |
| MGL | XPO1 | 2 | 61477849 | 61538626 | PCG | 0.393 | 4.48 | 3.83E-04 | 7.63E-02 |
| MSN.3 | EVL | 14 | 99971449 | 100144236 | PCG | -0.262 | -6.25 | 1.17E-05 | 6.62E-02 |
| MSN.3 | AL589843.1 | 9 | 96687050 | 96774318 | lncRNA | -0.349 | -5.84 | 2.54E-05 | 6.62E-02 |
| MSN.3 | POU2F2 | 19 | 42086110 | 42196585 | PCG | -0.433 | -5.70 | 3.26E-05 | 6.62E-02 |
| MSN.3 | TRMT9B | 8 | 12945642 | 13031503 | PCG | -0.329 | -5.45 | 5.32E-05 | 6.62E-02 |
| MSN.3 | HS3ST4 | 16 | 25691959 | 26137685 | PCG | -0.428 | -5.41 | 5.83E-05 | 6.62E-02 |
| MSN.3 | PBX1 | 1 | 164555584 | 164899296 | PCG | -0.233 | -5.36 | 6.33E-05 | 6.62E-02 |
| MSN.3 | PITPNM2 | 12 | 122983480 | 123150015 | PCG | -0.288 | -5.33 | 6.83E-05 | 6.62E-02 |
| MSN.3 | MPP6 | 7 | 24573268 | 24694193 | PCG | -0.284 | -5.23 | 8.26E-05 | 6.62E-02 |
| MSN.3 | AC025887.2 | 18 | 32470288 | 32745567 | lncRNA | -0.463 | -5.21 | 8.60E-05 | 6.62E-02 |
| MSN.3 | ANKRD36B | 2 | 97492663 | 97589965 | PCG | -0.369 | -5.18 | 9.13E-05 | 6.62E-02 |
| MSN.3 | PER1 | 17 | 8140472 | 8156506 | PCG | 0.226 | 4.93 | 1.52E-04 | 9.42E-02 |
| MSN.3 | NRSN1 | 6 | 24126186 | 24154900 | PCG | -0.231 | -4.87 | 1.71E-04 | 9.42E-02 |
| MSN.3 | RGN | X | 47078355 | 47093314 | PCG | -0.227 | -4.83 | 1.87E-04 | 9.42E-02 |
| MSN.3 | MYO9B | 19 | 17075781 | 17214537 | PCG | 0.298 | 4.82 | 1.89E-04 | 9.42E-02 |
| MSN.3 | PDE4A | 19 | 10416773 | 10469630 | PCG | -0.362 | -4.80 | 1.95E-04 | 9.42E-02 |
| MSN.3 | LINC01476 | 17 | 59422088 | 59526946 | lncRNA | -0.277 | -4.74 | 2.20E-04 | 9.97E-02 |
| OLI | MGAT5 | 2 | 134119983 | 134454621 | PCG | -0.317 | -5.51 | 4.76E-05 | 7.23E-02 |
| OLI | DICER1 | 14 | 95086228 | 95158010 | PCG | -0.219 | -5.07 | 1.14E-04 | 8.68E-02 |
| OPC | TMEM178B | 7 | 141074064 | 141480380 | PCG | -0.359 | -6.77 | 4.52E-06 | 1.09E-02 |

Note: ASC.1 is astrocyte group 1, MGL is microglia. MSN.3 is medium-sized spiny neuron group 3, OLI is oligodendrocytes and OPC is oligodendrocyte precursor cells. PCG is protein coding gene and lncRNA is long non-coding RNA. Parameter b is fixed effect regression coefficient for the gene in the mixed model and T its test statistic.

### Testing differences in cell-type composition

Alcohol use has been associated with phenomena such as disrupted neurogenesis(24, 25) and demyelination(26) that may affect the cell-type composition. Visual inspection of Figure S3 suggested that the clusters were similar for cases and controls. Using a mixed-model for binomial response data, we also found that nuclei from cases were not significantly more likely to be outliers than controls (odds ratio 1.17, P values 0.96). Finally, none of the 17 cell-types showed a significant difference in cell-type proportions between cases and controls (Table S5). Thus, little evidence was found for differences in cell-type composition in brain.

### Replication in pre-frontal cortex

We attempted replication using a s_n_-RNA-seq study in 3 alcohol dependent patients and 4 controls(4). This study involved pre-frontal cortex (PFC), which makes it less likely results replicate as there will be true differences between regions. Of the 136 replication tests (there were 6 reported PFC cell-types but P values for our 26 DEGs were occasionally missing), 13 yielded P < 0.05 (Tables S7 and S8). This is almost twice the 6.8 (=0.05×136) expected by chance (P value binomial test 0.019). Our MGL finding *CD53* replicated in PFC microglia. After a cell-type specific Bonferroni correction, our MSN.3 finding *MYO9B* remained significant in PFC astrocytes and *POU2F2* in PFC microglia.

### Pathways analyses

To study cell-type specific AUD disease processes, we first identified all genes expressed in a target cell-type (i.e., all genes passing QC). Next, Reactome(27) pathway analyses were performed in ConsensusPathDB(28) to study possible functions of that cell-type. Finally, we selected the pathways containing DEGs to identify cell-type functions relevant for AUD. This two-step approach allowed pathways analyses for all cell-types even if there were only a few significant findings.

The main MSN.3 pathway clusters (Figure 2, and Table S9) were related to axonal guidance and cell migration, metabolism of carbohydrates and lipids, the Rho family of small GTPases, circadian clocks, and neurotransmitter receptors and postsynaptic signal transmission. The main MGL clusters (Figure 2, and Table S10) were related to immune function, TGF-beta receptor signaling, and signaling by Rho GTPases. For the other cell-types we could not perform pathways analyses as the significant genes were not in any of the Reactome pathways.

**Figure 2.**
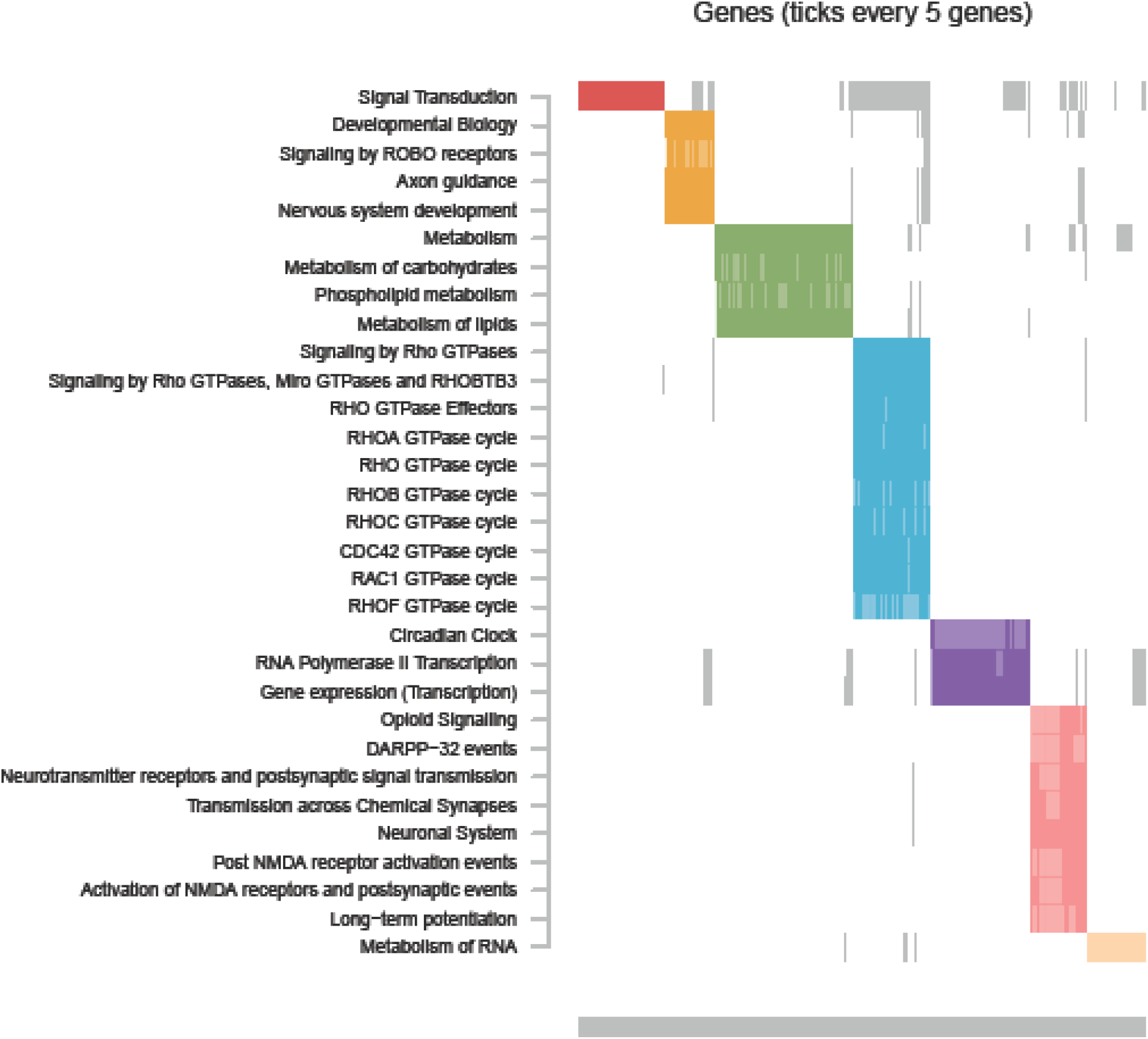
MSN.3: Significantly enriched pathways with AUD associated genes. The raster plot visualizes the clustering of pathways (y-axis) determined on the basis of their overlapping genes (x-axis). For clustering we used the Louvain Method for community detection(66). Only genes that were expressed in MSN.3 are plotted, rather than all possible pathway members. The solid-colored rectangles indicate genes assigned by the algorithm to the colored pathway cluster and that are members of the listed pathway, the transparent colored rectangles indicate genes assigned by the algorithm to the colored pathway cluster but that is not a member of the listed pathway, the grey rectangles indicate genes that are members of the listed pathway but that were assigned to a different pathway cluster. Complete pathway names, gene names, odds ratios, and P-values are presented in **Table S9**.

**Figure 3.**
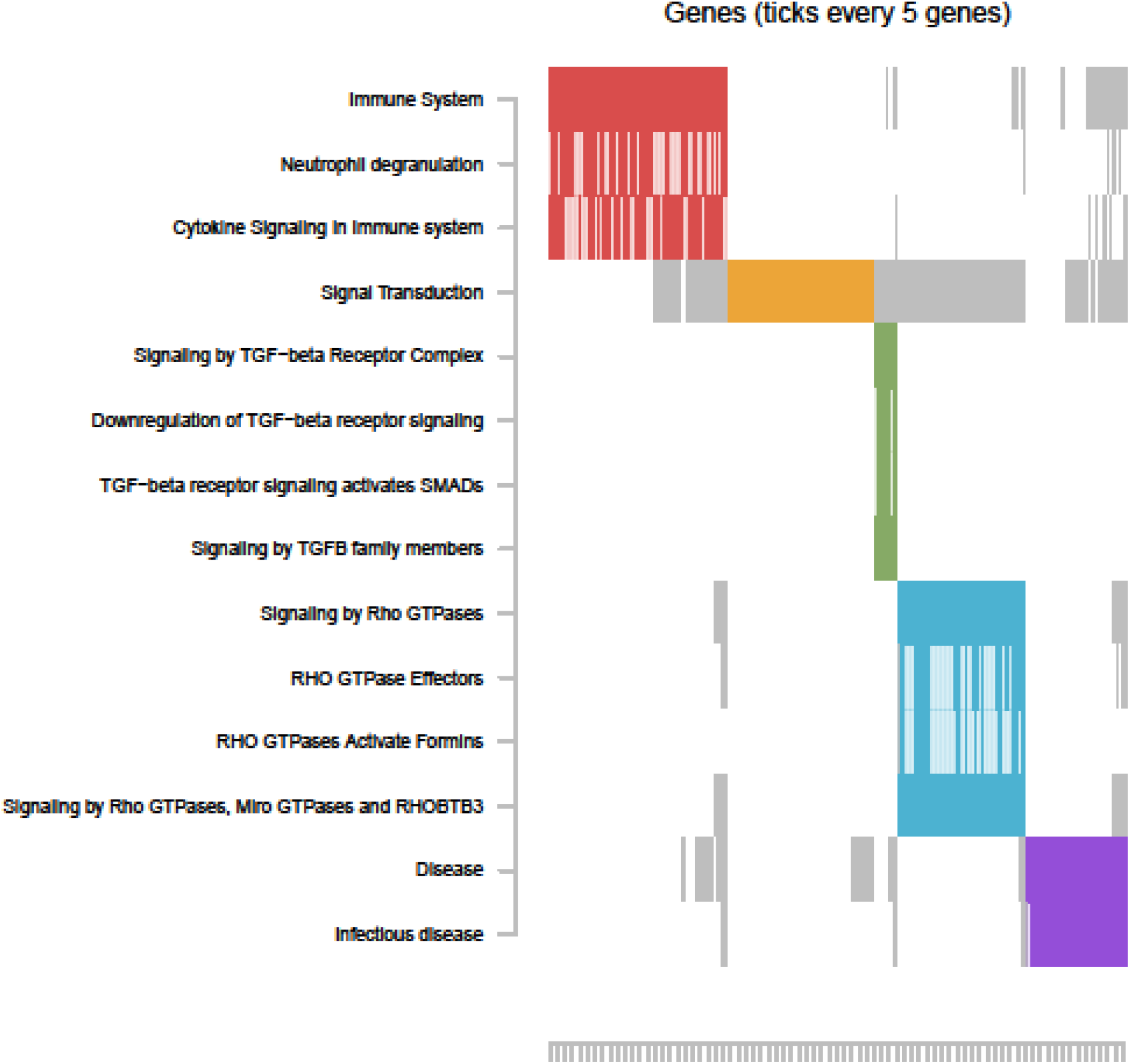
MGL: Significantly enriched pathways with AUD associated genes. See legend Figure 2 for description of the plot. Complete pathway names, gene names, odds ratios, and P-values are presented in **Table S10**.

## DISCUSSION

We performed a s_n_-RNA-seq study of AUD in the nucleus accumbens (NAc). The majority of transcriptome-wide significant genes involved a group of medium spiny neurons with both D1-type and D2-type dopamine receptors (MSN.3), microglia (MGL) and oligodendrocytes (OLI). A significantly higher number of findings than expected by chance replicated in a s_n_-RNA-seq study(4) of alcohol dependency in the pre-frontal cortex.

The top finding in MSN.3 was *PBX1*. A *PBX1* knock-out study in mice showed a decrease in the number of midbrain dopaminergic neurons(29), which is further supported by another study demonstrating that expression of *PBX1* is necessary for the survival of adult-generated neurons(30). One of the long-term effects of AUD is neurodegeneration which is characterized by the loss of function of neurons and, eventually, alcohol-induced apoptosis. Our study suggests a possible role of *PBX1* expressed mixed D1/D2 MSNs.

Circadian rhythm disturbances were implicated by our MSN.3 pathway analyses and a top finding in MSN.3 was *PER1. PER1* is a so-called CLOCK gene, which is part of the Period gene family that encode for components of circadian rhythm and has previously been linked to problematic alcohol use in humans(31, 32) and increased ethanol intake in mice(33, 34). Recent work has demonstrated that *PER1* is upregulated in the nucleus accumbens shell in a binge drinking model(33), which is congruent with our finding that *PER1* is upregulated in AUD cases. Alcohol disrupts circadian rhythms and circadian disruption is known to increase alcohol consumption(35). The dopaminergic receptor expression in NAc MSNs has been shown to be rhythmic and is thought to mediate the relationship between circadian rhythm and addiction.

Pathways analyses in MSN.3 further implicated the metabolism of carbohydrates and NMDA (*N*-methyl-d-aspartate receptor) receptors which are a key component of synaptic plasticity. The brain is the largest consumer of energy in the body. There is a growing body of evidence demonstrating that chronic alcohol consumption leads to alcohol-induced decreases in glucose metabolism(36). This decrease in glucose metabolism may reflect reductions in neuronal excitability, a shift toward alternative energy sources, or alcohol-induced neurotoxicity(37). Recent work has demonstrated that decreasing glucose metabolism is negatively associated with cortical thickness in heavy drinkers, suggesting a likely role for neurotoxicity in certain brain regions instead of a shift toward alternative energy sources(37). There is ample evidence that acute alcohol exposure induces inhibition of NMDA receptors in the nucleus accumbens while cycles of chronic exposure and withdrawal increase NMDA receptor function(38).

MGL are involved in immune response and neuroinflammation in the brain, which is also indicated by our pathway results and several of our top differentially expressed genes in this cluster such as *CD53* and *TRAF3*. Binge-like consumption of alcohol leads to MGL activation and heightened immune response in the brain of post-mortem AUD patients(39). MGL depletion (i.e., removal of MGL cells by genetic and pharmacological approaches(40), blunts the neuroinflammatory response after during alcohol withdrawal(41). *CD53*, a surface cell adhesion molecule, which is a known suppressor of inflammatory cytokine production(42) and may regulate other immune response pathways(42). *CD53* may mediate MGL migration(43) and has previously been linked to alcohol in rodents and humans(44-46). *TRAF3* is involved in immune response through its control of type 1 interferon production, which alcohol is known to inhibit(47). Another set of immune-related pathways implicated in MGL are related to the transforming growth factor beta (TGF-*β*) receptor signaling pathway. TGF-*β* is activated by an inflammatory event and, through the modulation of genes in the *SMAD* family, limits neuroinflammation resulting from the event(48). Alcohol has been shown to disrupt the TGF-*β* pathway in the brain, however, the focus has been primarily on the prefrontal cortex and hippocampus. In prefrontal cortex, mouse studies of chronic intermittent ethanol vapor exposure and voluntary consumption both showed decreased expression of genes in the TGF-*β* pathway in MGL(49, 50). Similarly, TGF-*β* mRNA levels in MGL were altered in the hippocampus of adolescent rats after several binge drinking sessions(51).

*MGAT5* and *DICER1* were significantly associated with AUD in OLI. OLI are the myelin-producing glial cells of the central nervous system. Myelin plays a critical role in neuronal communication by insulating the axon, enhancing the propagation of action potentials, and facilitating high-frequency firing(52). Alcohol is known to alter myelinating OLI, and myelinating OLI have been implicated in the establishment of addictive behavior and AUD(53). A study in mice showed that *MGAT5* reduces N-glycan branching, a process known to regulate oligodendrogenesis, primary myelination, and myelin repair(54). In humans, *MGAT5* has previously been linked to alcohol dependence in individuals with conduct disorder or suicide attempts(55). *DICER1* is also required for normal OLI differentiation and myelination(56). Another possible mechanism explaining the DICER1-AUD association involves β-catenin, a protein that among other functions regulates OLI development (57). *DICER1* has been implicated in stress and depression related phenotypes, both risk factors for AUD, in humans and mice(58). A study in mice found that β-catenin in D2-type medium spiny neurons in the nucleus accumbens mediates such affects by activating a network that includes *DICER1* as a key target gene(59).

Almost all detected pathways were cell-type specific. The exception involved Rho GTPases that were implicated in MSN.3 and MGL. In comparison to other drugs of abuse, the impact of alcohol on Rho GTPase activity has not received much attention. However, a few studies have linked genes in the Rho GTPase pathway to alcohol consumption and sensitivity(60, 61). In response to extracellular signals, Rho GTPases can induce coordinated changes in the organization of the actin cytoskeleton(62). In neurons, these cytoskeletal elements give the cell dynamic structure and are involved in a myriad of processes, including dendritic spine growth and morphology. The latter processes seem to be especially relevant to drug abuse, as ‘inappropriate’ learning of strongly reinforcing cues is a hallmark in the development of addiction and changes in the number of dendritic spines have long been known to occur after repeated exposure to drugs of abuse(63). For MGL, the promotion of Rho GTPases is linked to neuroinflammation which suggests the potential for Rho GTPases to mediate the AUD-linked proliferation of MGL activation to increased neuroinflammatory response(64).

Several limitations of the present study should be mentioned. First, statistical power in s_n_RNA-seq studies depends on a variety of factors such as the number of donors, number of nuclei, the number of reads and the amount of debris remaining after sample preparation. Although our study compares favorably to existing s_n_RNA-seq studies in terms of these power determining factors, it is likely that we may have detected only the genes with large effects and that a larger number of donors are needed to detect more modest effect sizes(65). Furthermore, the use of a larger number of donors may avoid atypical donors creating idiosyncrasies in the data and thereby improve the generalizability of findings. In our study, we aimed to investigate the expression profile in the NAc of AUD cases and controls. However, there were some indications that our tissue samples were predominantly from the NAc shell. Explicitly sampling tissue from both the shell and core as well as expanding the study to include multiple brain regions will generate a more complete picture AUD pathogenic processes of the brain. Finally, s_n_RNA-seq studies in post-mortem brains are correlational in nature meaning that it is unclear whether the discovered pathways changed as a result of alcohol use or contributed to the susceptibility to AUD. Functional follow-up studies will be needed to shed light on the causal direction of effects.

We performed the first s_n_-RNA-seq study of AUD in the NAc, a region that is key due to its central role in the mesolimbic reward pathway. The functions of the detected genes and pathways for each cell-type were consistent with the functions of those cell-types reported in the literature. Part of our findings involved genes and pathways previously associated with AUD where we could advance current knowledge by linking altered gene expression to one of three specific cell-types. The genes and pathways that had not previously been linked to alcohol had high face validity in the broader context of addiction and suggested new avenues for studying AUD. Some of our findings seemed particularly robust. For example, the altered *CD53* expression in MGL replicated in a s_n_-RNA-seq study of a different brain region, and was previously implicated in a bulk gene expression studies in rodents(44) and studies of DNA sequence variants(45) and methylation(46) in humans. The identification of the specific cell-types from which these association signals originated will facilitate designing the proper follow-up experiments and, eventually, may result in new and targeted clinical interventions.

## Data Availability

Data will become publicly available as soon as the article is accepted for publication

## ACKNOWLEDGEMENTS

This work was supported by grant 5R01AA026057 (PI Clark) and a NARSAD Young Investigator Grant from the Brain & Behavior Research Foundation (PI Clark). Post-mortem tissue samples were obtained from the Human Brain Collection Core through the NIH NeuroBioBank. We would like to thank Dr. Dayne Mayfield for providing summary statistics from their s_n_-RNA-seq study in pre-frontal cortex for replication.

## CONFLICT OF INTEREST

The authors declare that they have no conflict of interest.

## AVAILABILITY OF DATA AND MATERIALS

Data are available from GEO under accession number # and will be publicly available as soon as the article is accepted for publication

## Notes

### Competing Interest Statement

The authors have declared no competing interest.

### Author Declarations

Texas A&M University IRB, Reference number 097974, Date November 04 2019: Ruling "The Institution determined that the proposed activity is not research involving human subjects as defined by DHHS and FDA regulations." Virginia Commonwealth University IRB, Reference number HM20019402, Date May 18 2020 Ruling "VCU IRB review or approval is not required before you proceed with your involvement this project". As this research involves deceased individuals is not human subjects research according to 45 CFR 46.102(f).

