## Supplemental material for "A single-nucleus transcriptomics study of alcohol use disorder in the nucleus accumbens"

**SUPPLEMENTAL MATERIAL FOR THE PAPER:** A single-nucleus transcriptomics study of alcohol use disorder in the nucleus accumbens by van den Oord et al.

### METHODS

#### Nuclei isolation

Collecting the nuclei: For each sample, 15 to 30 mg of frozen tissue was dounced with 20 strokes and a loose pestle in 1ml of homogniztion buffer with RNasin Plus, followed by 10 strokes with a tight pestle. The homogenization buffer (see Table S1 for details). Two ml of the same buffer was added, the homogenate was passed through a 70-μm cell strainer and 2 ml of homogenization buffer without RNasin Plus was added. Next, the homogenate was passed through a 30-μm cell strainer, repeated twice, each time using a fresh strainer and collection tub. The volume of the collected homogenate was brought up to 10 ml with homogenization buffer without RNasin Plus and centrifugated in 4°C for 10 minutes at 900 x g to pellet the nuclei.

Removing myelin Three ml of blocking buffer (see Table S1 for details) was added and the pellet was resuspended by pipetting. Next, 30 ul of myelin removal beads (Miltenyi Biotech, 130-096-733) was added and incubated in 4ºC for 15 minutes followed by adding 3 ml of blocking buffer and centrifugation in 4°C for 5 minutes at 300 x g to pellet the nuclei and beads. The supernatant was removed and the pellet was resuspended by pipetting in 3 ml new blocking buffer. The resuspension was divided into equal amounts (1ml /tube) across three 1.5 ml LowBind tubes and placed on a microtube magnet (DynaMag-2 Magnet, Invitrogen) in 4ºC for 15 minutes. Following the incubation, from each tube, 800 ul of the supernatant was passed through a 35 um Cell strainer Snap Cap and collected in a Falcon Round-bottom test tube (5ml) that was centrifugated in 4°C for 5 minutes at 300 x g with a swing bucket. All but 100 ul of the supernatant was removed, 300 ul of blocking buffer was added and the nuclei suspension was passed a second time through the Snap Cap strainer.

Quantifying the nuclei: Quantification of the nuclei suspension was performed using a Countess II FL Hemocytometer and default settings.

#### Single nucleus RNA sequencing

We have used the Chromium platform from 10X Genomics in combination with the Chromium Next GEM Single Cell 3' Reagent Kits v3.1 (Dual Index) for partitioning of 1,000 nuclei per sample, cDNA conversion and library preparation. All lab-technical procedures were performed following the vendor's recommendations(1). As described above, quantifications determined by the Countess II FL Hemocytometer (described above) was used to identify the amount of nuclei suspension to load on the Chromium to achieve the desired number of nuclei. However, it should be noted, for four samples, an error with the Countess settings resulted in that a larger number of nuclei were partitioned than originally intended. Following library construction each library was sequenced on a NovaSeq 6000 using a 200 cycle reagent kits with the second read that contains the cDNA sequence being 102bp

#### Table S1. Stock solutions and buffers

| Item: | Components: | Volume or mass | Final Concentration: |
|---|---|---|---|
| | Stock Solutions (Store at +4 ºC) | | |
| 20 ml 10% BSA | BSA | 2 g | 10% |
| | Nuclease-free water | 20 ml | |
| 20 ml 10% Triton X-100 | Triton X-100 | 2 ml | 10% |
| | Nuclease-free water | 18 ml | |
| 250 ml Nuclei Isolation Media (NIM) pH 8.0 | Nuclease-free water | 221.675 ml | |
| | 1 M Tris Buffer (pH 8.0) | 2.5 ml | 10 mM |

| | | | |
|---|---|---|---|
| | Sucrose | 21.45 g | 250 mM |
| | 2 M KCl | 3.125 ml | 25 mM |
| | 1 M $MgCl_2$ | 1.25 ml | 5 mM |
| 50X Protease Inhibitor Cocktail | 50X Protease Inhibitor Cocktail Stock Bottle | | 50X |
| | 100% EtOH | 1ml | |
| Required Fresh Solutions (Store on ice or in a 4°C fridge until ready to use) | | | |
| 25 ml Blocking Buffer | Nuclease-free water | 19.875 ml | |
| | 10X PBS, pH 7.4 | 2.5 ml | 1X |
| | 10% BSA | 2.5 ml | 1% |
| | 40 U/ul RNasin Plus | 125 ul | 0.5%, 0.2X |
| 25 ml Homogenization Buffer | NIM pH 8.0 | 24.225 ml | |
| | 100 mM DTT | 25 ul | 0.1 mM |
| | 50X Protease Inhibitor Cocktail | 500 ul | 1X |
| | 40 U/ul RNasin Plus | # ul | # |
| | 10% Triton X-100 | 250 ul | 0.10% |

#### Alignment

For sake of convenience we will use the term gene, but all transcripts with a poly-A tail are captured (i.e., the vast majority of transcripts except, for instance, ribosomal RNAs generated by RNA polymerase I and III, other small RNAs generated by RNA polymerase III, and replication-dependent histone mRNAs, and a few long non-coding RNAs synthesized by RNA polymerase II.(2)). To account for characteristics that are specific to libraries from the Chromium platform, the cellranger(3) software was used for de-multiplexing of nuclei, aligning the reads, and creating a matrix of unique molecular identified (UMI) counts (i.e., the number of unique molecules for each gene detected in each nucleus).

Chromium $s_n$RNA-seq data primarily yields reads derived from mature spliced RNA (mRNA) and mapping to exonic regions. Additionally, it may capture unspliced pre-mRNA transcripts that can generate intronic reads(4-6). As nuclei contain a relatively large fraction of pre-mRNA molecules and such molecules are particularly abundant in brain tissue(7), it is recommended to count intronic reads as well(8). We therefore aligned reads to GRCh38 with the “include-introns” option in cellranger that uses a gene transfer format (GTF) file that allows for intronic alignments.

Table S2 provides sequencing statistics. In summary, we obtained an average 283,165,911 reads per sample of which 94.8% mapped to the genome, with 92.7% RNA read bases with Q-score >= 30 and where 81.5% of reads had nucleus-associated barcodes.

Table S2. Sequencing statistics (separate file)

#### Quality control

<u>Sample QC:</u> Figure S1 shows so-called barcode rank plots that plot the total barcode count against the rank of each barcode where the highest ranks have the largest totals. Barcodes for nuclei should have significantly more counts associated with them than the barcodes of background “noise”. The cellranger(3) software uses this principle to estimate the number of nuclei for each assay.

Figure S1 Barcode rank plots

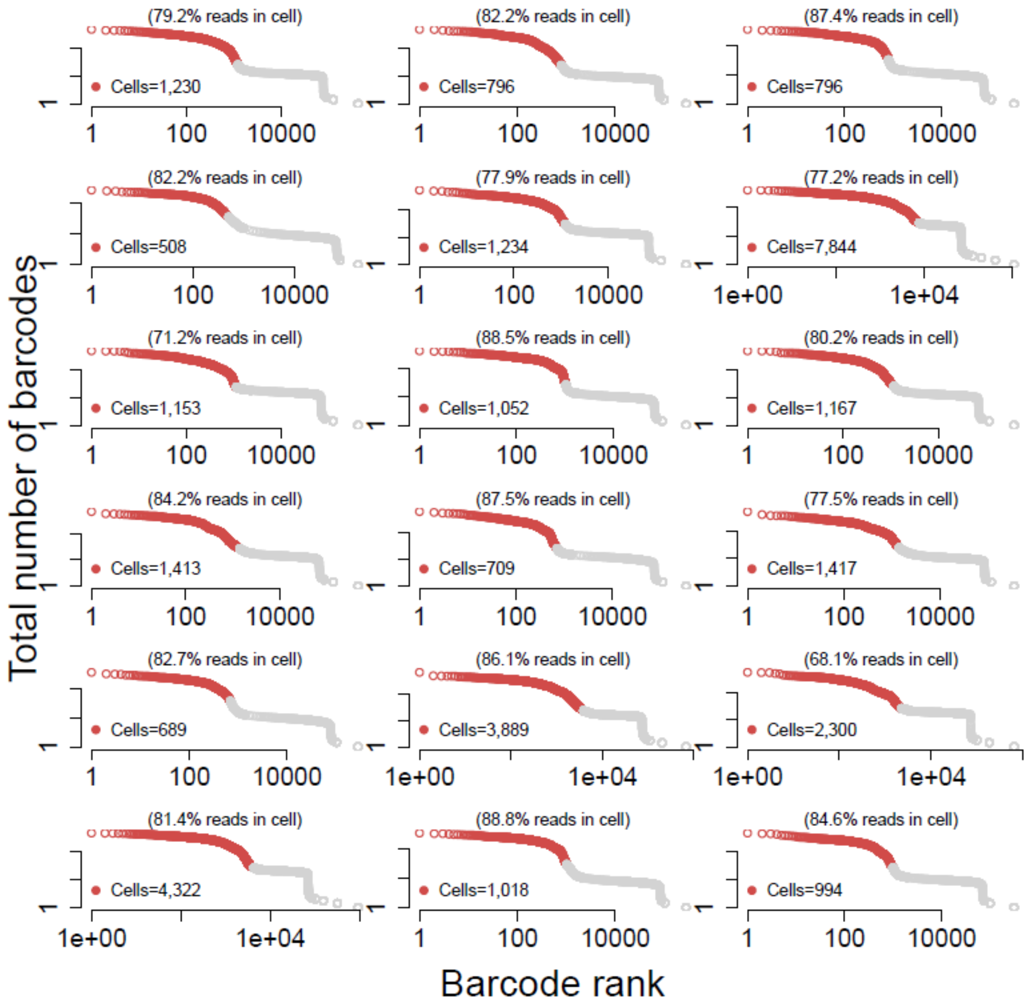

Nuclei QC: Across the 18 samples there were an estimated 32,531 nuclei. Low-quality nuclei or empty droplets are likely to have few genes expressed and a small number of UMI counts, whereas nuclei multiplets are likely to have a high gene and UMI count as they capture expression levels of multiple nuclei. We removed 1,492 nuclei with fewer than 600 genes and more than 10,000 genes. These thresholds were chosen as they defined the extreme values of the distribution of number of genes per nucleus (Figure S2A, we used the log base 10 as this transformation was also used prior to performing cluster and association analyses). Next, we eliminated 130 nuclei with UMI counts < 650 and with UMI counts > 100,000 (Figure S2B). Finally, we removed 165 nuclei with more than 5% of reads mapping to ribosomal genes as that may be an artifact stemming from sample preparation. This left 32,531 - (1,492+130+165) = 30,744 nuclei (94.5%)

Figure S2. Distribution and QC threshold for number of genes and UMI counts per nucleus

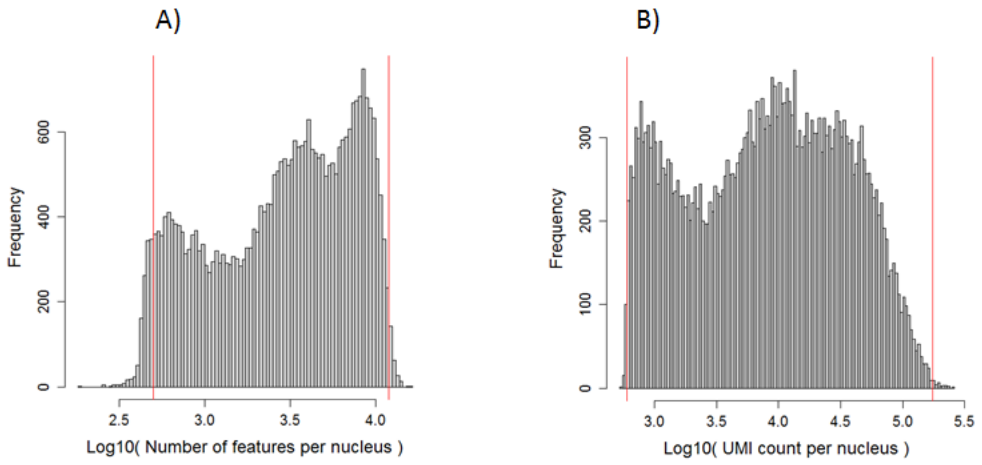

Gene QC: Our data comprised 36,601 genes. For the cluster analyses the 2,000 most highly variable genes were selected (see next section). Prior to the association analyses we removed low abundant genes to avoid inaccurate test statistics caused by analyzing sparse data. As expression levels of genes can vary substantially between nuclei, this QC step was performed for each cell-type separately rather than across all nuclei. This avoids, for example, that genes that are highly expressed in only one cell-type are eliminated because of low expression levels for all the other cell-types. Specifically, for each cell-type separately we eliminated (i) all genes observed in less than 10% of the nuclei, and (ii) all genes with a mean abundance less than 2 across all nuclei of that cell-type. The number of genes retained after this QC was on average 4,287 genes across per cell-types.

#### Clustering

To identify cell-type clusters we used Seurat(9). UMI count data were log-normalized to obtain more normal distributions and reduce effects of possible outliers. Next, to give equal weight and avoid that highly-expressed genes dominate the cluster analyses, the data was scaled to have a mean expression across nuclei of zero and a variance of one. Limiting analyses to genes that exhibit high nucleus-to-nucleus variation (i.e., highly expressed in some nuclei and lowly expressed in others) typically improves results from cluster analyses(10). The 2,000 most highly variable genes were selected. Subject-level variation (e.g., due to demographic variables) and confounders (e.g., cDNA yield, percentage of reads aligned) plus lab technical artefacts may obscure the separation of clusters. We therefore regressed out (i) “dummy” variables that indicated the 18 subjects, and (ii) Chromium batch and (ii) the QC measures discussed above (i.e., log 10 number of genes per nucleus, log 10 UMI counts per nucleus, and % of reads mapping to ribosomal genes). The log-normalized, scaled and residualized data were then submitted to a principal components analysis (PCA). Next, we constructed a K-nearest neighbor graph based on the Euclidean distance of the space defined by the 15 PCs that explained most of the variation in the data and further refined the edge weights between any two nuclei based on the shared overlap in their local neighborhoods. Finally, t-Distributed Stochastic Neighbor Embedding)(11) (t-SNE) was used to visualize the cell-type clusters in a two-dimensional space. t-SNE is a non-linear dimensionality reduction tool that can preserve local structure in low dimensional space. This means that points that are close to one another in the higher-dimensional PCA space, will tend to be close to one another in the low dimension (this is, for example, not the case for PCA or other linear dimensionality reduction techniques that subsequently may provide a less clear picture of the clusters).

##### Outliers

As a final QC step we eliminated nuclei that were outliers with respect for the cell-type they were assigned to. We also For this purpose, we calculated for each nucleus an outlier score that was the mean of the absolute differences between all the feature means for that nucleus and the corresponding feature means across all nuclei of that cell-type. Next, we calculated the median absolute deviation (MAD) of these outlier scores and identified outliers as nuclei with absolute( outlier score - median( outlier score)) / MAD( outlier score) > 3. We use the MAD rather than standard deviations to define outlier as the MAD is the more robust measure(12).

#### RESULTS

##### Cell-type identification and labeling

###### Table S3. Cell-type cluster markers (separate file)

###### Table S4. Labeling with known expression markers (separate file)

To study possible differences between cases and controls we performed an integrated analysis in Seurat(9). This was achieved by first identifying matching nuclei between the two groups called 'anchors' followed by clustering all nuclei in the shared space defined by the anchors(13). Figure S3 shows that nuclei for cases (blue) and controls (red) overlap with the identified clusters (Figure 3A) and that cell-type clusters cluster were very similar between cases and controls (Figure 3B).

Figure S3. Cell-type clusters for AUD cases and controls.

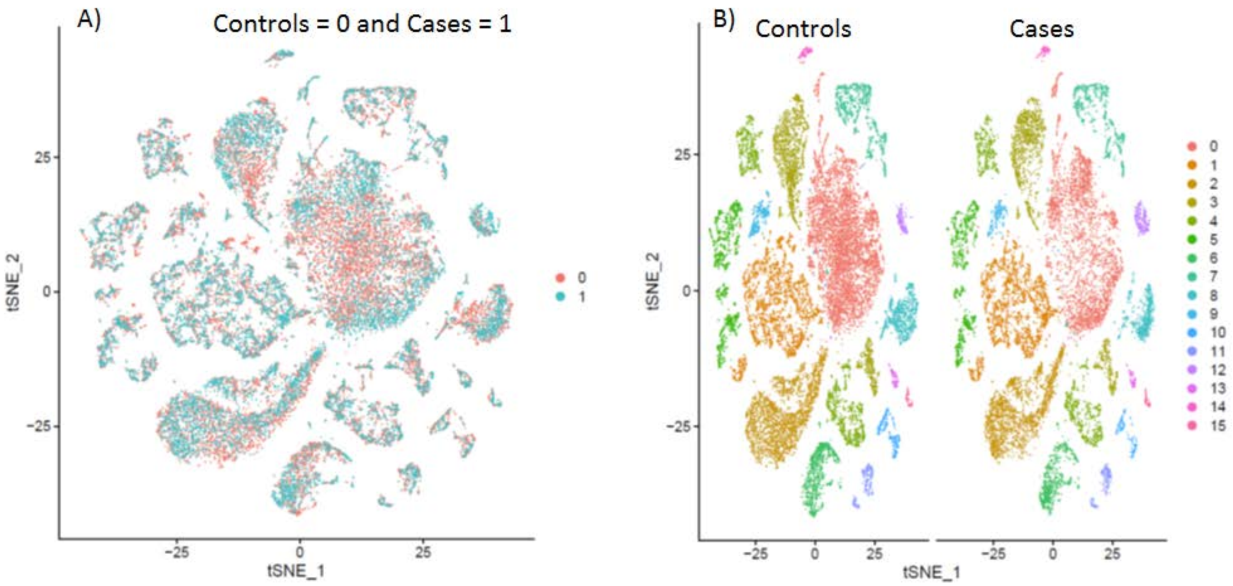

###### Table S5. Case-control cell-type proportions (separate file)

#### Identifying differentially expressed genes (DEGs)

##### Table S6. Significant suggestive association findings (separate file)

Table S7 gives a summary of the replication Brenner at al.(14) that involves A final study involved nuclei from the pre-frontal cortex of 4 alcohol dependent patients and 3 controls(14). The actual P values and other replication statistics are given in Table S8

Table S7 Summary replication using results from the study by Brenner at al.

| Cell | Gene | ASC | EX | IN | MGL | OLI | OPC |
|---|---|---|---|---|---|---|---|
| ASC.1 | CCDC18.AS1 | n-- | n-- | n-- | n-- | n-- | n-- |
| MGL | CEP85L | yn- | yn- | yn- | yn- | yn- | yn- |
| MGL | SUMF1 | yyn | yyn | yn- | yn- | yn- | yn- |
| MGL | TRAF3 | yn- | yn- | yn- | yn- | yn- | yn- |
| MGL | CD53 | yn- | yn- | yn- | yyn | n-- | yn- |
| MGL | RAPH1 | yyn | yn- | yn- | yn- | yn- | yn- |
| MGL | XPO1 | yn- | yn- | yn- | yn- | yn- | yn- |
| MSN.3 | EVL | yyn | yn- | yn- | yyn | yn- | yn- |
| MSN.3 | AL589843.1 | yn- | yn- | yn- | yn- | yn- | yn- |
| MSN.3 | POU2F2 | yn- | yn- | yn- | yyy | yn- | yn- |
| MSN.3 | TRMT9B | yn- | yn- | yn- | yn- | yn- | yyn |
| MSN.3 | HS3ST4 | yn- | yn- | yn- | yn- | yn- | yn- |
| MSN.3 | PBX1 | yn- | yn- | yn- | yn- | yyn | yn- |
| MSN.3 | PITPNM2 | yn- | yn- | yn- | yn- | yn- | yn- |
| MSN.3 | MPP6 | yn- | yn- | yn- | yn- | yyn | yn- |
| MSN.3 | AC025887.2 | yn- | yn- | yn- | yn- | yn- | yn- |
| MSN.3 | ANKRD36B | yyn | yn- | yn- | yn- | yn- | yn- |
| MSN.3 | PER1 | n-- | n-- | n-- | n-- | n-- | n-- |
| MSN.3 | NRSN1 | yn- | yn- | yn- | yn- | yn- | yn- |
| MSN.3 | RGN | yn- | yn- | yn- | n-- | yn- | yn- |
| MSN.3 | MYO9B | yyy | yn- | yn- | yn- | yn- | yn- |
| MSN.3 | PDE4A | n-- | n-- | n-- | n-- | n-- | n-- |
| MSN.3 | LINC01476 | yn- | yn- | yn- | yn- | yn- | yn- |
| OLI | MGAT5 | yn- | yn- | yn- | yn- | yn- | yn- |
| OLI | DICER1 | yyn | yn- | yn- | yn- | yn- | yn- |
| OPC | TMEM178B | yn- | yn- | yn- | yn- | yn- | yn- |

Note: Rows indicate findings significant in our study after FDR control at 0.1 (for cell-type labels see Table 2 main text). Columns refer to the cell-types tested in the study by Brenner at al. where ASC is astrocytes, EX is excitatory neurons, IN is GABAergic inhibitory neurons, MGL is microglia, OLI is oligodendrocytes and OPC is oligodendrocyte precursor cells. The entries are a three letter string indicating whether in the in the study by Brenner et al. the 1) test results were available, 2) results were significant allowing for a Type I error of 0.05, and 3) results were significant after a cell-type specific Bonferroni correction allowing for a family-wise error rate of 0.05. Values in the string are y is yes, n is no, and – is not applicable.

##### Table S8. Replication statistics (separate file)

#### Pathway analyses

Table S9. MSN.3 Reactome pathway results (separate file)

Table S10. MGL Reactome pathway results (separate file)
